# Home-based and remote exercise testing in chronic respiratory disease, during the COVID-19 pandemic and beyond: a rapid review

**DOI:** 10.1101/2020.07.15.20154930

**Authors:** Anne E Holland, Carla Malaguti, Mariana Hoffman, Aroub Lahham, Angela T Burge, Leona Dowman, Anthony K May, Janet Bondarenko, Marnie Graco, Gabriella Tikellis, Joanna Y. T. Lee, Narelle S Cox

**Affiliations:** Department of Allergy, Immunology and Respiratory Medicine, Monash University; Department of Physiotherapy, Alfred Health; Institute for Breathing and Sleep, Australia.; Department of Cardiorespiratory and Skeletal muscle, Federal University of Juiz de Fora, Brazil.; Department of Allergy, Immunology and Respiratory Medicine, Monash University, Australia.; Department of Allergy, Immunology and Respiratory Medicine, Monash University; Department of Physiotherapy, Austin Health; Institute for Breathing and Sleep, Australia.; Department of Allergy, Immunology and Respiratory Medicine, Monash University; School of Exercise and Nutrition Sciences, Institute for Physical Activity and Nutrition (IPAN), Deakin University, Australia.; Department of Physiotherapy, Alfred Health, Australia.; Allied Health, Alfred Health; Institute for Breathing and Sleep, Australia.; Department of Allergy, Immunology and Respiratory Medicine, Monash University; Institute for Breathing and Sleep, Australia.

**Author notes:** Corresponding Author: Anne E Holland, Department of Allergy, Immunology and Respiratory Medicine, Monash University Commercial Rd Melbourne 3004.

**Keywords:** Exercise Test, Lung Diseases, Rehabilitation, Home Care Services, Telemedicine

## Abstract

**Objectives:** To identify exercise tests that are suitable for home-based or remote administration in people with chronic lung disease.

**Methods:** Rapid review of studies that reported home-based or remote administration of an exercise test in people with chronic lung disease, and studies reporting their clinimetric properties.

**Results:** 84 studies were included. Tests used at home were the 6-minute walk test (6MWT, 2 studies), sit-to-stand tests (STS, 5 studies), Timed Up and Go (TUG, 4 studies) and step tests (2 studies). Exercise tests administered remotely were the 6MWT (2 studies) and step test (1 study). Compared to centre-based testing the 6MWT distance was similar when performed outdoors but shorter when performed at home (2 studies). The STS, TUG and step tests were feasible, reliable (intra-class correlation coefficients >0.80), valid (concurrent and known groups validity) and moderately responsive to pulmonary rehabilitation (medium effect sizes). These tests elicited less desaturation than the 6MWT, and validated methods to prescribe exercise were not reported.

**Discussion:** The STS, step and TUG tests can be performed at home, but do not accurately document desaturation with walking or allow exercise prescription. Patients at risk of desaturation should be prioritised for centre-based exercise testing when this is available.

## Introduction

As a result of the COVID-19 pandemic, many pulmonary rehabilitation programs have transitioned rapidly to remote delivery models.^1, 2^ Whilst studies have shown it is possible to deliver exercise training, physical activity counselling, education and self-management training remotely, all existing clinical trials have included an in-person exercise test prior to program commencement, to assess safety of exercise (e.g. degree of oxyhaemoglobin desaturation) and enable accurate exercise prescription.^3, 4^ During the COVID-19 pandemic centre-based or in-person assessments of exercise capacity are not able to be performed in most centres. As a result, some pulmonary rehabilitation programs have commenced exercise testing at home, using tests with minimal space requirements such as sit-to-stand (STS) or step tests, and with or without remote monitoring of oxyhaemoglobin saturation (SpO_2_) and heart rate. Other programs are not conducting any exercise testing prior to commencing patients on pulmonary rehabilitation programs at home. It is not clear which of our current tests of functional exercise capacity are suitable for home and / or remote administration.

The research questions for this rapid review were:

1. Which functional exercise tests have been conducted in the home setting in people with chronic lung disease?
2. Which functional exercise tests have been conducted remotely in people with chronic lung disease?
3. What are the clinimetric properties of tests that have been conducted at home or remotely, including feasibility, reliability, validity and responsiveness to pulmonary rehabilitation?
4. Can these functional exercise tests be used to assess safety (particularly oxyhaemoglobin saturation) and prescribe exercise intensity, either in-person or remotely?

## Methods

The protocol was registered on PROSPERO (CRD42020182375) on 27^th^ April 2020. Inclusion criteria are presented in Box 1.

### Types of studies

We included any study that reported conducting an exercise test at home or remotely in people with chronic respiratory disease. We also included studies conducted in any setting that report use of tests that were being conducted at home in people with chronic respiratory disease during the COVID-19 pandemic, specifically step tests and sit-to-stand (STS) tests.^1^. These studies were included in order to report on their clinimetric properties (quality of measurement instruments e.g. reproducibility) and clinical properties (e.g. ability to detect desaturation and prescribe exercise). We did not include studies that reported the clinimetric properties of the 6-minute walk test (6MWT) in a centre-based setting, as these have been reported in detail in a previous systematic review.^5^

We did not include case studies. Review articles were not included, but we reviewed their reference lists for studies that met our inclusion criteria. Otherwise there were no restrictions on study design. We included studies investigating clinimetric properties, descriptive studies and studies where the test was used to evaluate the effects of an intervention. Only studies published in English were included.

### Participants

We included studies in which participants had any chronic lung disease including (but not limited to) chronic obstructive pulmonary disease (COPD), interstitial lung disease (ILD), asthma, cystic fibrosis (CF), bronchiectasis or pulmonary hypertension. We did not exclude studies based on age, gender or physiological status of participants. We excluded studies that focused on participants who were mechanically ventilated.

### Search methods for identification of studies

As this was a rapid review designed to respond to the emerging COVID-19 pandemic, we elected to search MEDLINE from year 2000 to 25^th^ April 2020. The search strategy for MEDLINE is in supplementary Table 1. One author reviewed the title and abstract of the identified studies to determine their inclusion.

#### Data extraction and management

One author conducted data extraction using a standardised template, with random checks on accuracy by a second reviewer. The following information was extracted:

- Methods of study (date/title of study, aim of study, study design, primary outcome, other outcomes)
- Participants (diagnosis, age, sex, disease severity, inclusion criteria, exclusion criteria, method of recruitment of participants)
- Intervention (if applicable, description of the intervention)
- Exercise test - name, details of protocol (if provided), location of test (home, centre, other) and monitoring (in person, remote, none), variables monitored
- Outcomes pre/post intervention data where applicable, details of clinimetric properties if applicable
- Details of any physiological monitoring, including but not limited to pulse oximetry
- Whether the results of the test were used to prescribe exercise and if so, the methods used.

#### Assessment of risk of bias

We considered risk of bias according to study design and methods of analysis, and this was documented in the data extraction form. As this was a rapid review we did not conduct a formal assessment using a risk of bias tool.

#### Outcomes

The main outcomes of interest were the number of reports of home or remote administration of each exercise test. Additional outcomes were patient variables monitored for each test (e.g. SpO_2_, heart rate, symptoms, blood pressure); methods used to prescribe exercise training intensity; and clinimetric properties for each test - feasibility, reliability, validity and responsiveness, using the metrics reported by the authors.

#### Data synthesis

A narrative synthesis was performed for each exercise test separately. For each exercise test we reported whether it had been performed at home or with remote monitoring, including the number of reports. Patient variables monitored for each test (e.g. SpO_2_, heart rate, symptoms, blood pressure) were reported descriptively. Any methods used to prescribe exercise training intensity were reported descriptively.

We reported clinimetric properties for each test, from all studies where these are reported, not just those performed at home. We reported feasibility (eg number of participants who could perform the test), reliability (e.g. intra-class correlation coefficient (ICC)), validity (e.g. correlation with gold standard exercise tests) and responsiveness to pulmonary rehabilitation (e.g mean changes pre/post rehabilitation and measures of variability). Where possible we calculated an effect size to describe responsiveness.

We had intended to examine outcomes separately by subgroups with different lung diseases (e.g. COPD, ILD), but there were insufficient data for diseases other than COPD, so these analyses were not performed.

## Results

The MEDLINE search identified 3778 studies (excluding duplicates) of which 3654 were excluded based on title and abstract. Of the 128 full text papers screened, 84 were included (85 reports). This included five studies examining the 6MWT^6–10^, 39 studies examining STS tests,^11–49^ 35 studies examining step tests^18, 23, 49–81^ and 17 studies examining the Timed Up and Go (TUG).^16, 18, 23, 32, 41, 48, 49,82–91^

Ten studies examined more than one test, including three that examined three tests.^18,23,49^ The PRISMA diagram is in Figure 1 and study characteristics are in Supplementary Tables 2-5. An overall summary of the review findings is in Figure 2.

**Figure 1.**
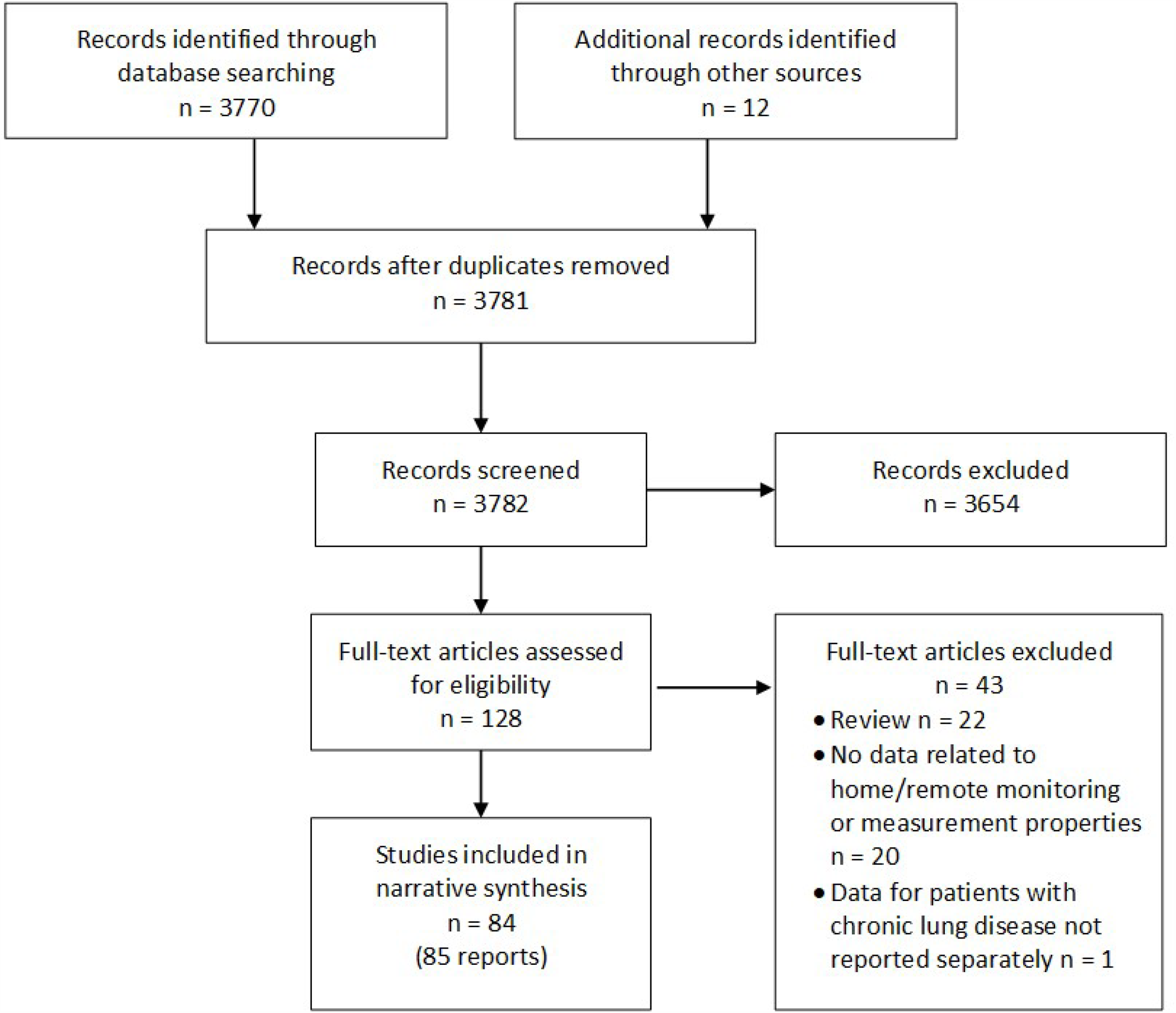
Study selection.

**Figure 2.**
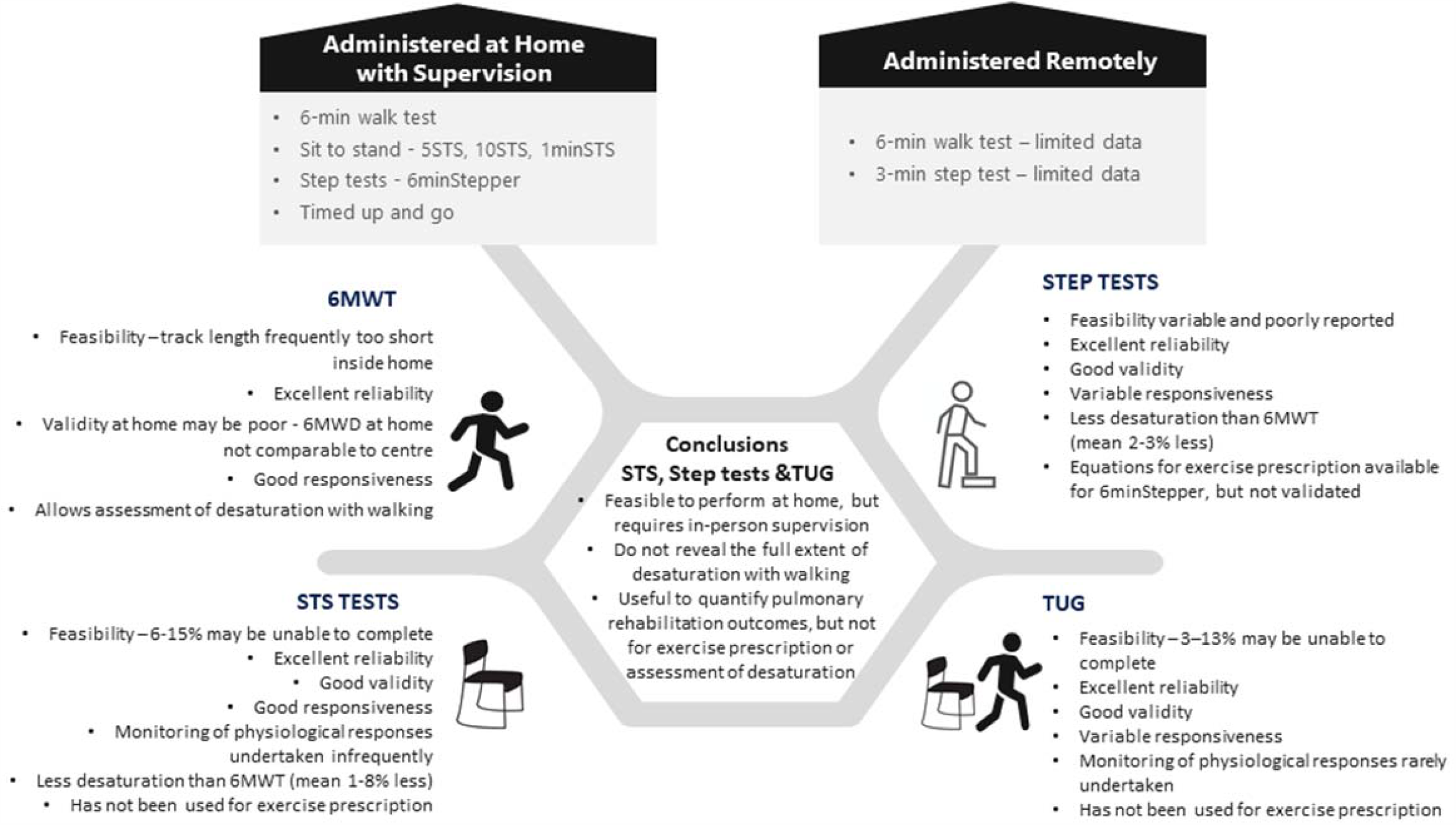
Summary of review findings. 6MWD = distance walked on 6-minute walk test, 6MWT = 6-minute walk test, STS = sit to stand, TUG = Timed Up and Go

### Main outcome – home and remote use

Exercise tests that have been used at home in people with chronic lung disease were the 6MWT (2 studies),^6, 7^ five times STS (5STS, 2 studies),^33, 41^ ten times STS (10STS, 1 study, 2 reports),^18, 23^ 1-minute STS (1minSTS, 1 study),^49^ 6-minute stepper test (6minStepper, 2 studies, 3 reports), ^18, 23, 49^ and TUG.^18, 23, 41, 49, 91^ Exercise tests administered remotely were the 3-minute step test (3MST)^58^ and 6MWT.^8, 9^

### 6-minute walk test

#### Home

One randomised crossover trial (RXT) compared home and centre-based 6MWTs^7^ and one RXT compared an outdoors to a centre-based 6MWT.^6^ Both included people with moderate to severe COPD. The centre-based 6-minute walk distance was significantly longer than the distance recorded at home^7^ (Table 1) with a mean difference that exceeded the minimal important difference of 30 metres.^92^ The 6MWT track lengths were shorter at home (mean 17 metres) compared to the centre (30 metres) and 42% of tests were conducted indoors. Comparison of indoor vs outdoors 6MWT (conducted on a flat sidewalk), both using a 30-metre track, showed no difference in the distance walked(Table 1).^6^

**Table 1.** Difference between centre-based and home or remote test administration.

| Test | Study | Comparison | Difference |
| --- | --- | --- | --- |
| 6MWT | Holland 2015<sup>7</sup> | Centre vs home | 6MWD mean 30.4 metres longer at the centre (95%CI 0.4 to 63.2 metres) |
|  | Brooks 2003<sup>6</sup> | Indoor vs outdoors | 6MWD mean (SD) 394 (86) vs 398 (84) metres, p=0.4 |
|  | Juen 2014<sup>8</sup> | App vs in-person | 6MWD MD 0.3m (95%CI -73 to 72 metres)<br>App absolute error for 6MWD 5.87% |
|  | Juen 2015<sup>9</sup> | App vs in-person | App error for 6MWD 3.78% |
| 3MST | Cox 2013<sup>58</sup> | Remote supervision vs in-person | Nadir SpO<sub>2</sub> MD 0.2% (LOA - 3.4 to 3.6%)<br>Rate of perceived exertion MD 0.5 points (LOA - 1.1 to 2.1 points)<br>Pulse rate MD - 0.6 beats/min (LOA -11.3 to 10.1 beats/min). |

3MST - 3-minute step test, 6MWD - 6-minute walk distance, 6MWT - 6-minute walk test, 95%CI - confidence interval, LOA - limits of agreement, MD - mean difference, SD - standard deviation, SpO<sub>2</sub>- oxyhaemoglobin saturation.

#### Remote

Two studies by the same group aimed to validate two different phone apps for remote monitoring of the 6MWT in people with chronic respiratory conditions (mostly COPD and asthma).^8, 9^ Both apps recorded the 6-minute walk distance using accelerometry, and one also provided voice and vibrating instructions.^8^ Both apps included monitoring by pulse oximetry, however these data were not reported. The 6-minute walk distance measured by the apps was similar to that measured by the researchers in person (Table 1).

#### Feasibility

One study in participants with COPD reported that 58% of tests were conducted outdoors because a track of sufficient length was not available inside the home. ^7^

#### Clinimetric properties

Home-based 6-minute walk distance was highly reliable when performed twice on the same day, with ICCs ≥ 0.99. ^7^ Intra-rater reliability was high for both outdoor and indoor tests (ICCs 0.97 and 0.99 respectively).^7^

#### Safety assessment

All studies reported monitoring the 6MWT using pulse oximetry and three also used symptom scales for dyspnoea and perceived exertion.^6, 7, 10^

#### Exercise prescription

One study used the 6MWT for exercise prescription in 39 people with COPD.^10^ Walking exercise was prescribed at 80% of the average speed walked on the 6MWT. This exercise prescription was well tolerated over 10 minutes of walking, generally achieving more than 60% of peak oxygen uptake (VO_2_) with a steady state by the 4^th^ minute.

### Sit-to-stand tests

#### Six different STS tests were used (Table S2)

These were the five times sit to stand test (5STS, 14 studies), where the time taken to stand up and sit down five times from a standard height chair is recorded; the ten times sit to stand test (10STS, 2 studies) using a similar protocol; the 30-second sit to stand test (30secSTS, 9 studies) where the number of sit-to-stand repetitions in 30 seconds is recorded; the 1-minute sit-to-stand test (1minSTS, 13 studies) as well as small numbers of studies using 2-minute tests (2minSTS, 1 study) and 3-minute tests (3minSTS, 2 studies).

#### Home

Tests used at home were the 5STS,^33, 41^ 10STS, ^18, 23^ and the 1minSTS.^49^ Participants (n=381) had COPD, some were using home oxygen therapy^49^ and some were recovering from an acute exacerbation.^33^ All home testing involved in-person supervision from a researcher or clinician.

#### Remote

No studies reported remote administration or monitoring of a STS test.

#### Feasibility

In a study of patients with stable COPD (n=475), 15% of participants were unable to complete the 5STS.^26^ Those who were unable to complete the test were significantly older (mean (SD) 73(10) vs 68(10) years), had higher levels of chronic dyspnoea (Medical Research Council scale 4.1(1.0) vs 3.3(1.1) points), lower quadriceps maximal voluntary contraction (44(13) vs 60(17)%predicted) and lower incremental shuttle walk distance (84(66) vs 224 (126) metres). A study comparing the 5STS to the 30secSTS in 128 people with moderate to severe COPD reported that all participants could complete the 5STS but 7% could not complete two trials of the 30secSTS.^44^ One additional trial reported that 3 of 50 participants with COPD (6%) could not complete any repetitions of the 30secSTS.^25^ Of those participants who felt it was strenuous to undergo a STS (69%), most (93%) found the 30secSTS more strenuous than the 5STS.^44^ In a clinical trial of inpatient pulmonary rehabilitation including 60 participants with moderate to severe COPD, all could complete both the 30secSTS and the 1minSTS.^43^ No feasibility data were reported for the 10STS, 2minSTS or 3minSTS.

#### Clinimetric properties

Reliability, validity and responsiveness of STS tests are in Table 2. Test-retest reliability was high for the 5STS, 30secSTS and 1minSTS. The 5STS, 30secSTS and 1minSTS had moderate to strong correlations with other measures of exercise capacity, with higher values for the 1minSTS than the other tests. There were moderate correlations with quadriceps strength and weak correlations with daily life physical activity. Predictive validity was demonstrated only for the 1minSTS, with lower values predicting increased mortality at 2 and 5 years.^20, 35^ Responsiveness to pulmonary rehabilitation was evident for 5STS, 30secSTS and 1minSTS, with moderate to large effect sizes.

**Table 2.**
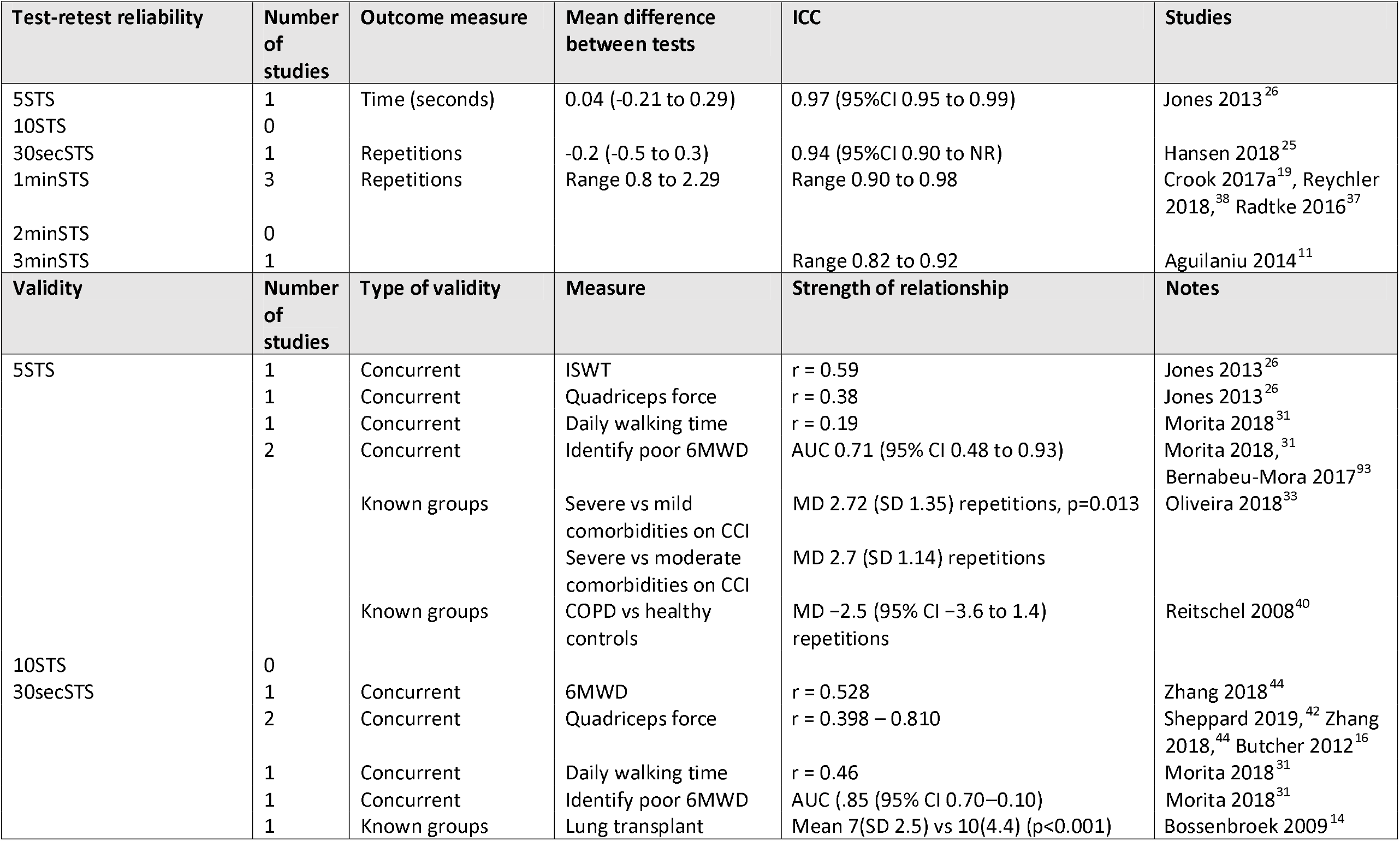
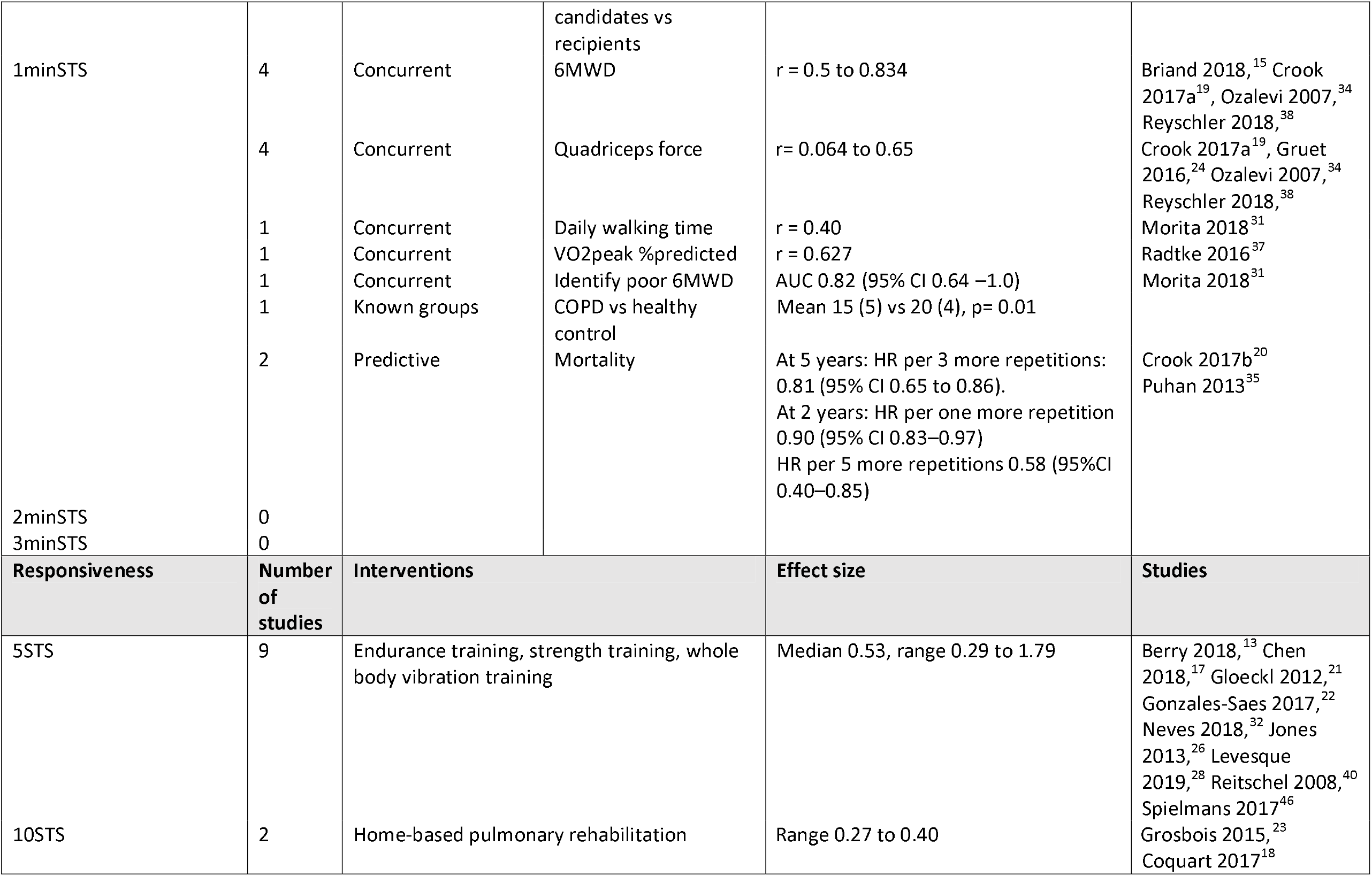
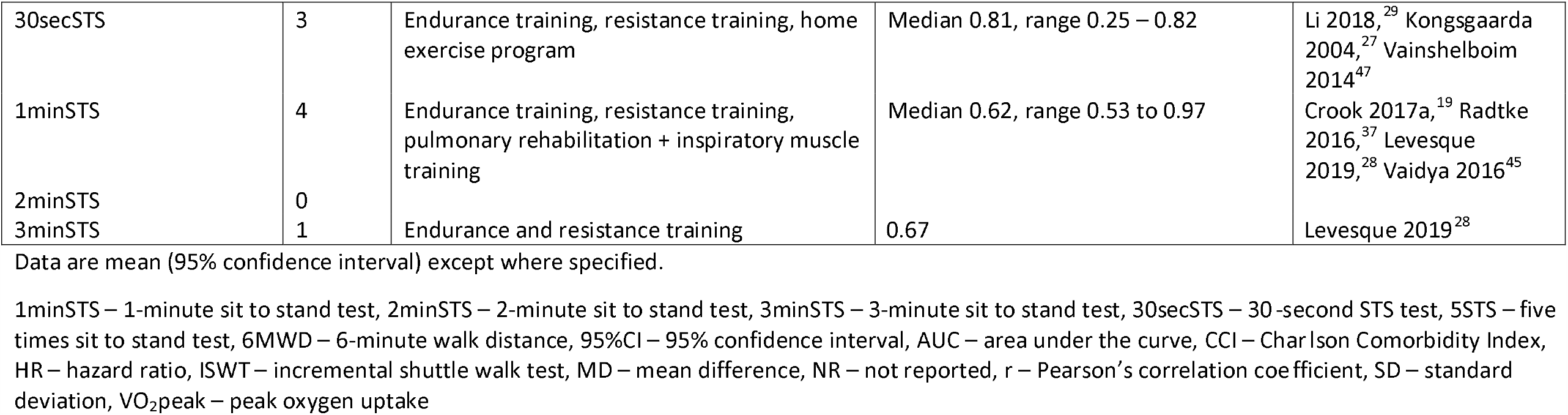
Clinimetric properties of sit-to-stand tests.

#### Safety assessment

Most studies did not report using any monitoring during the STS test (24 / 40 studies, 60%, Table S2).

A comparison of three STS tests in people with COPD found significantly greater desaturation on the 1minSTS than the 30secSTS or 5STS (mean −3(SD 4) vs −1(2) and −1(2) respectively).^31^ Greater desaturation on 1minSTS than 30secSTS was reported in a second study in COPD (mean −2.6 (2) vs 2(1.8).^43^ The 1minSTS also gave rise to significantly greater increases in heart rate than the 30secSTS or 5STS (mean 22(13) vs 16 (10) and 7(7)) and higher fatigue scores (median 2 vs 0.5 vs 0).^31^ Dyspnoea scores on 1minSTS did not differ from the 30secSTS but were significantly greater than 5STS (median 2.5 vs 1 vs 0) with a similar pattern of findings for systolic blood pressure (median 30 vs 20 vs 0 mmHg).^31^

In comparison to the 6MWT and cardiopulmonary exercise test (CPET), the 1minSTS provoked less oxyhaemoglobin desaturation and a smaller rise in heart rate (Table 3). The VO_2_peak was also significantly lower during 1minSTS than during the CPET (median 1.68 [IQR 1.38, 2.29] vs 1.25 [1.03, 1.86]).^36^ Symptom scores for dyspnoea and fatigue were variable, with some studies reporting that they were similar across the tests,^24, 38^ higher on CPET than 1minSTS,^36^ higher on 6MWT than 1 minSTS,^34^ or higher on 1minSTS than 6MWT.^15^

**Table 3.** Fall in oxyhaemoglobin saturation and rise in heart rate on 1-minute sit-to-stand test compared to conventional exerci se tests.

| Study | Patient group | Oxyhaemoglobin desaturation (SpO<sub>2</sub>%) |  |  | Maximum heart rate |  |  |
| --- | --- | --- | --- | --- | --- | --- | --- |
|  |  | 1minSTS | 6MWT | CPET | 1STS | 6MWT | CPET |
| Briand 2018<sup>15</sup> | ILD | 92 (5) | 90 (7) |  | 112 (17) | 112 (16) |  |
| Crook 2017a<sup>19</sup> | COPD | 90 (3) | 86 (6) |  | 107 (11) | 107 (15) |  |
| Gruet 2016<sup>24</sup> | CF | -4 (3) | -5 (4) | -7 (5) | 131 (18) | 141 (16) | 171 (14) |
| Ozalevi 2007<sup>34</sup> | COPD | 0 (1) | -3 (3) |  | 98 (22) | 110 (20) |  |
| Radtke 2017<sup>36</sup> | CF | -6 [-3 to -9] |  | -9 [6 to 11] | 154 [148 to 159] |  | 169 [166 to 178] |
| Reyschler 2018<sup>38</sup> | COPD | -1 (3) | -8 (5) |  | 14 (10) | 20 (15) |  |

Data are mean (SD) or median [interquartile range]. Data are decrease in SpO<sub>2</sub> from baseline, with the exception of Briand et al and Crook et al, which are nadir SpO<sub>2</sub>.
1minSTS – 1-minute sit-to-stand test; 6MWT – 6-minute walk test; CF – cystic fibrosis, COPD – chronic obstructive pulmonary disease, CPET – cardiopulmonary exercise test; ILD – interstitial lung disease

Exercise prescription: No studies used any of the STS tests for exercise prescription.

### Step tests

#### Five different step tests were used (Table 3)

6-minute stepper test (6MStepper)(15 studies), using a hydraulic stepper; a 3-minute step test (3MST)(9 studies), most at a fixed cadence (7 studies); incremental step tests (5 studies), where the stepping rate increases regularly throughout the test, using either the Chester protocol (4 studies) or a version modified for patients with lung disease (modified incremental step test, MIST, 3 studies); a step oximetry test (4 studies) involving either stepping on and off a single step 15 times (3 studies) or for as long as possible (1 study); and a 6- minute step test on a single step at a free cadence (2 studies).

#### Home

Two studies (3 reports) used the 6MStepper to assess exercise capacity before and after a rehabilitation program at home.^18, 23, 49^ These tests used a hydraulic stepper with in-person supervision in the home. Participants (n=337) had moderate to severe COPD and some were using long-term oxygen therapy.

#### Remote

One study compared a remotely supervised 3MST to a 3MST monitored in person in 10 adults with CF and moderate lung disease.^58^ Remote supervision took place via videoconferencing and included measures of SpO_2_ and pulse rate via pulse oximetry, with the monitor visible to the health professional via videoconferencing. Measures of dyspnoea and perceived exertion were also collected. There was good agreement between the directly supervised and remotely supervised tests for nadir SpO_2_, pulse rate and rate of perceived exertion (Table 1). Nine of ten participants indicated no preference for in-person or remote supervision, with one participant preferring in-person supervision.

#### Feasibility

Feasibility varied across the different step tests. One study reported that in patients with bronchiectasis the Chester Step Test was not as well tolerated as the MIST, which starts at a lower cadence and increases more slowly.^56^ The Chester Step Test was stopped more frequently than the MIST by the examiner (58% vs 41% of tests), either because the participant could not maintain the cadence, or due to desaturation.^56^ In contrast the entire 3MST at fixed cadence was completed by 97 of 101 adults with CF.^67^ One study reported that all participants (n=84 with ILD) could complete the 6minStepper test,^63^ however people using supplemental oxygen were not included. Some studies excluded participants with orthopaedic problems that would have prevented them undertaking the test,^75^ making it difficult to assess the feasibility of tests across the population of people with chronic lung disease.

#### Clinimetric properties

Reliability, validity and responsiveness of step tests are in Table 4. The 6minStepper, MIST and Chester step tests demonstrated good test-retest reliability, with limited data for other tests. Although the ICC for the 6minStepper was high (0.94) the second test recorded up to 42 steps more than the first test, due to warming of the hydraulic jacks in the stepper device.^54, 57^ There was some evidence of criterion validity for all tests, with moderately strong correlations to other important measures such as 6-minute walk distance or physical activity in daily life. Data for responsiveness to pulmonary rehabilitation was only available for the 6minStepper and 3MST (free cadence), with variable effect sizes.

**Table 4.**
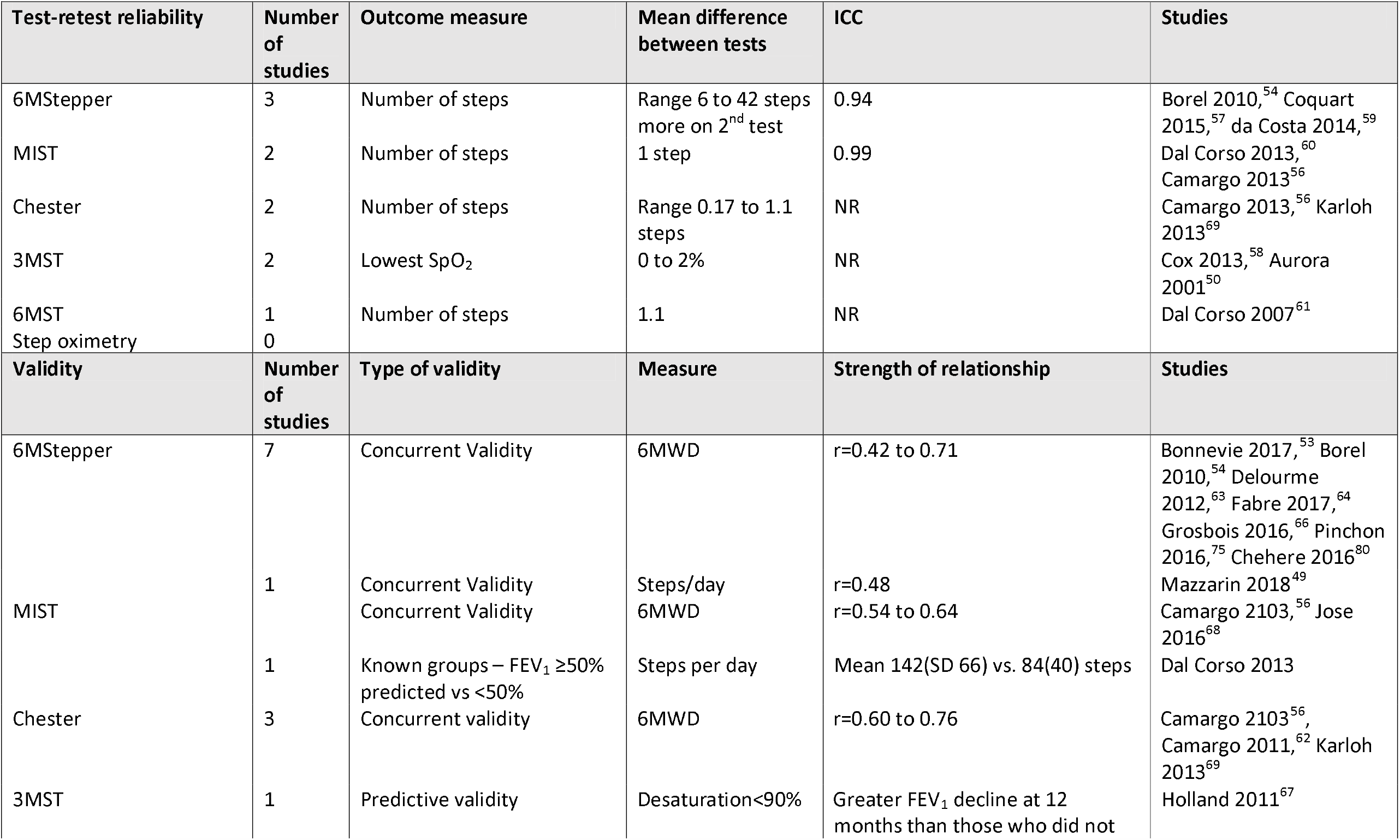
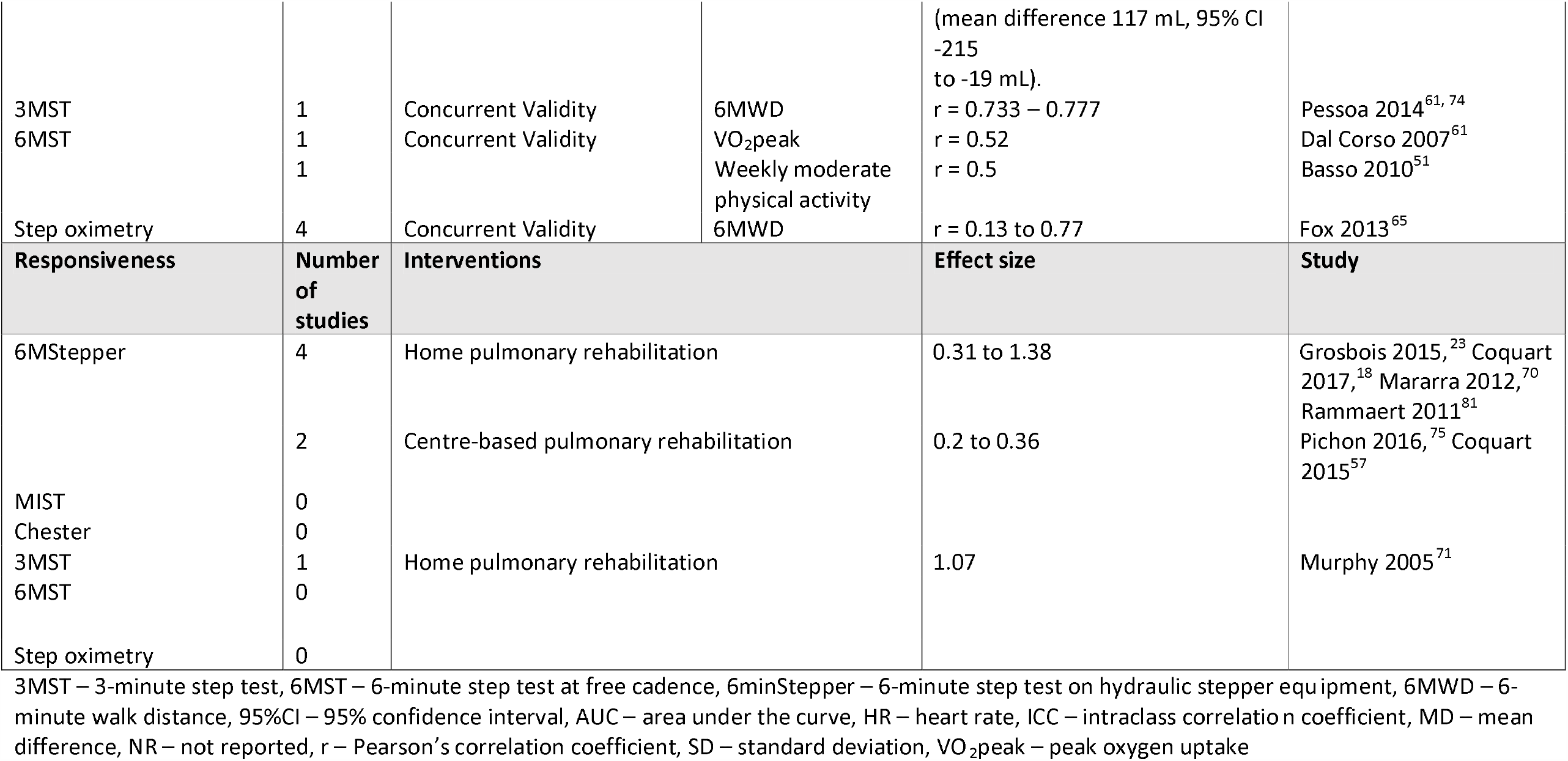
Clinimetric properties of step tests.

#### Safety assessment

All studies reported monitoring step tests with pulse oximetry and most also used symptom scales for dyspnoea and perceived exertion (Table S3). Several studies reported that the degree of desaturation was less on the 6minStepper than on 6MWT (SpO_2_ 2.3 to 3% more desaturation on 6MWT, 4 studies).^63, 70, 75, 80^ Desaturation on the 6MST with free cadence was not different to 6MWT^51^ or CPET.^61^ In contrast, an incremental step test (MIST) resulted in greater desaturation than a CPET (−7(5)% vs −3(3)%), but with similar rise in heart rate and similar symptoms.^60^ A 6MST with free cadence caused a greater rise in heart rate and more lower limb fatigue than a 6MWT, ^51^ with similar findings for the 6minStepper.^63^

#### Exercise prescription

Three studies of the 6minStepper had developed equations for exercise prescription. Two studies generated reference equations for prescribing aerobic training based on heart rate during the 6minStepper, but the equations were not validated.^53, 64^ and there were no reports of their use to set training intensity in pulmonary rehabilitation programs. A third study developed reference equations for prescription of resistance training and compared actual vs predicted training load (70% of 1 repetition maximum (1RM)).^52^ The mean difference was 30kg, and the authors concluded this difference was not clinically acceptable and the prediction equation should not be used as a substitute for a 1RM measure.

### Timed Up and Go

#### Home

The TUG was administered at home in 4 studies (5 reports),^18, 23, 41, 49, 91^ where it was used to evaluate the effects of a home pulmonary rehabilitation program^18, 23, 41, 49^ or to evaluate change over 12 months.^91^ Participants (n=381) had moderate to severe COPD (FEV_1_ %predicted mean 27 to 42%) and some were using home oxygen therapy.^49^ All home testing involved in-person supervision from a researcher or clinician.

#### Remote

No studies reported remote administration or monitoring of the TUG.

#### Feasibility

Two studies reported excluding participants who could not perform the TUG (13% and 3% of those recruited).^88, 89^

#### Clinimetric properties

Reliability, validity and responsiveness of the TUG are in Table 5. Test-retest reliability was high. Concurrent validity was demonstrated by moderate to strong relationships between TUG time and other measures of exercise capacity (6-minute walk distance, peak work, peak VO_2_) and peak quadriceps force, although one study reported no relationship between leg press and TUG time (data not reported).^48^ The TUG time was longer in fallers than non-fallers, and in oxygen users vs non-oxygen users.^82, 84, 85^ Responsiveness varied, with effect sizes ranging from small to large, and the minimal detectable change (95%) ranging from 14 to 33.5%.

**Table 5.**
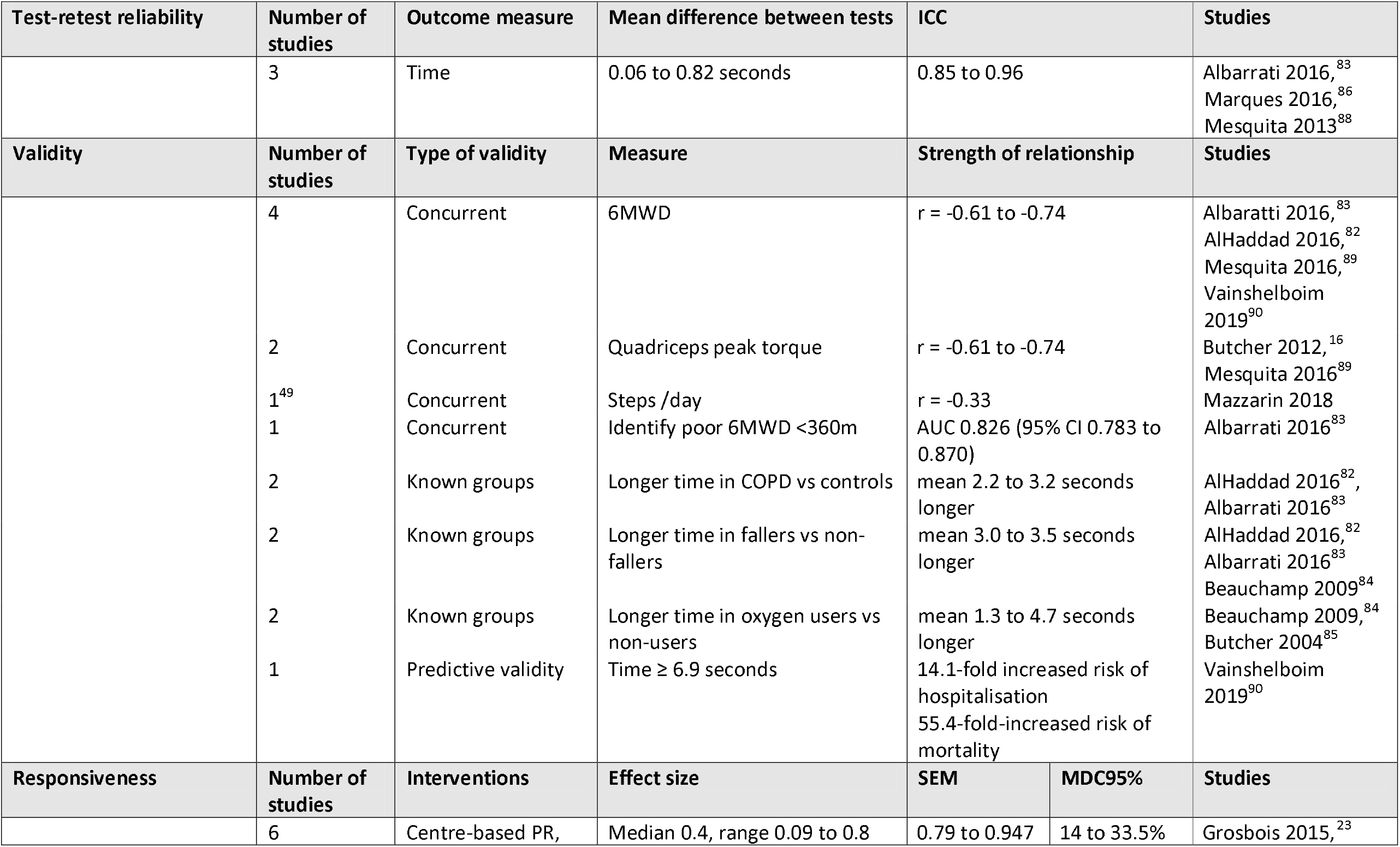
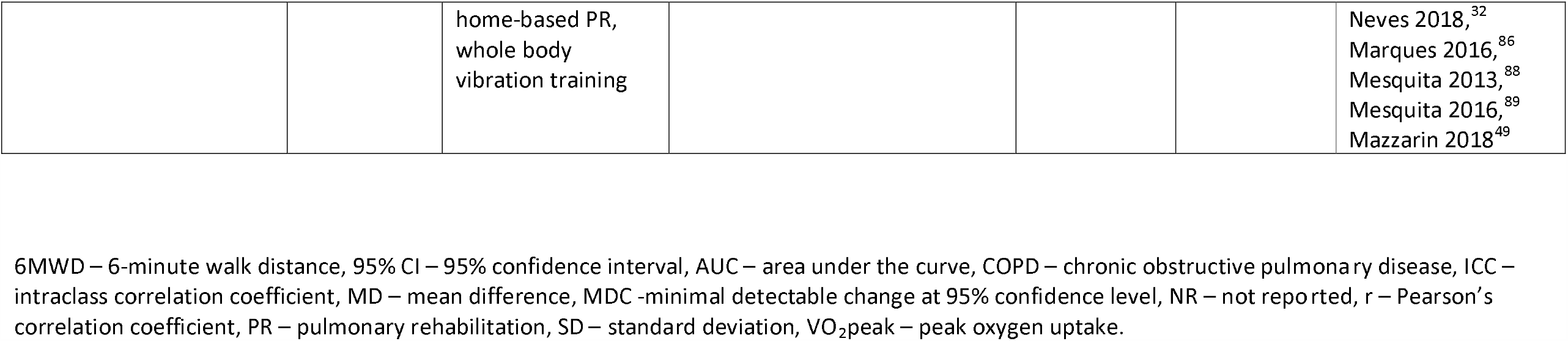
Clinimetric properties of Timed Up and Go.

#### Safety assessment

Only one out of 16 studies (6%) reported any monitoring of physiological variables during the TUG (Table S4).

#### Exercise prescription

No studies used the TUG to prescribe exercise.

## Discussion

This rapid review identified a range of exercise tests that have been used at home with supervision in people with chronic lung disease (6MWT, STS, 6minStepper and TUG) and a more limited range of tests that have been administered remotely (6MWT, 3MST). Administration of the 6MWT at home may be limited by short track lengths inside the house, although outdoors administration may provide a valid alternative where this is possible. The STS, step tests and TUG are feasible to perform in the home environment but do not reveal the full extent of desaturation with walking. These tests are useful to quantify improvements in physical function with home-based pulmonary rehabilitation but a gap remains in exercise prescription. Consideration should be given to identifying patients at risk of desaturation in whom centre-based exercise testing should be prioritised when local circumstances allow this to be performed safely.

This rapid review addresses an important challenge for pulmonary rehabilitation clinicians during the COVID-19 pandemic. Whilst delivery of pulmonary rehabilitation programs at home is feasible^3, 4^ and international bodies are advocating for remote delivery,^1, 2^ assessment of exercise capacity remains a key gap for many services. This review identifies a number of simple exercise tests that can be performed at home with supervision, when social distancing restrictions allow. These tests allow quantification of pulmonary rehabilitation outcomes, which is particularly important to evaluate in the context of a rapidly changing model of care. The small number of studies on remote administration of the 6MWT and 3MST provides some evidence that this approach would be feasible in selected patients (eg those not at risk of falls), but more data are required. Whilst the 6minStepper has been used to prescribe exercise in a small number of studies, reliability of this test may be limited by the equipment required, which appears to require a variable warm up period for the hydraulic jacks.^54, 57^ Outdoors administration of a 6-min walk test may be possible in some settings,^6^ depending on local weather and physical environment, which would allow both assessment of desaturation and prescription of exercise. This approach may prove more acceptable to some patients than an in-home or centre-based test, allowing social distancing to be better maintained.

Limitations to this review relate to both the body of evidence and the review process. The included studies often included a small number of participants and used a wide variety of testing protocols, which limited data synthesis. Feasibility of the tests was poorly documented and key patient groups were often excluded from studies (e.g. those using oxygen therapy or those who could not perform the test. Clinimetric properties of tests were rarely assessed in the home setting, but given the nature of the tests (STS, step and TUG) and the use of face-to-face supervision, these seem unlikely to vary substantially from those properties documented in centre-based testing. A small number of studies were available for patient groups other than COPD. A rapid review process was selected to ensure we could quickly address the immediate challenge facing the pulmonary rehabilitation community, but inherent limitations to this process must be acknowledged, including searching a single electronic database (Medline), a single author undertaking study selection, and a single author performing data extraction with accuracy checks on a random sample by a second reviewer. As this was a rapid review we did not perform a formal quality assessment, although data extraction included risk of bias related to study design and analysis, which was considered during data synthesis.

In conclusion, pulmonary rehabilitation clinicians can confidently perform STS, step and TUG tests at home in people with chronic lung disease, where in-person supervision is possible. Remote supervision may also be possible in selected patients, although few data are available. These in-home tests are useful to quantify the outcomes of home-based pulmonary rehabilitation, but do not reveal the full extent of desaturation on exercise, and validated methods to prescribe exercise intensity are not available. Consideration should be given to identifying patients at risk of desaturation in whom centre-based exercise testing should be prioritised, when local circumstances allow this to be performed safely.

### Source(s) of support

CM is partially supported by the Conselho Nacional de Desenvolvimento Científico e Tecnológico (CNPq) (process number: 200042/2019-0), and Coordenação de Aperfeiçoamento de Pessoal de Nível Superi – Brazil (CAPES) – Finance Code 001. NSC holds a National Health and Medical Research Council (NHMRC) Early Career Fellowship (GNT 1119970).

## Data Availability

All data included in this review are from studies in the public domain.

## Box 1.

**Inclusion criteria**

### Design

- Any study that reported conducting an exercise test at home or remotely in people with chronic respiratory disease
- Any study reporting clinimetric properties of these tests
- Case studies, review articles and articles not in English were excluded

### Participants

- People with chronic lung disease including (but not limited to) chronic obstructive pulmonary disease (COPD), interstitial lung disease (ILD), asthma, cystic fibrosis, bronchiectasis or pulmonary hypertension

### Intervention

- No intervention was necessary for inclusion

### Outcome measures

- Main outcome was the number of reports of home or remote administration of each exercise test
- Clinimetric properties for each test - feasibility, reliability, validity and responsiveness
- Patient variables monitored for each test (e.g. SpO_2_, heart rate, symptoms, blood pressure)
- Methods used to prescribe exercise training intensity

## Notes

### Competing Interest Statement

The authors have declared no competing interest.

### Author Declarations

This was a rapid review of previously published studies and no ethical approvals were required.

## References

1. Gardiner L, Graham L, Harvey-Dunstan T, et al. Pulmonary Rehabilitation Remote Assessment. British Thoracic Society. https://brit-thoracic.org.uk/about-us/covid-19-information-for-the-respiratory-community/. Accessed 15th May, 2020.

2. Garvey C, Holland AE and Corn J. Pulmonary Rehabilitation Resources in a Complex and Rapidly Changing World. https://www.thoracic.org/members/assemblies/assemblies/pr/resources/pr-resources-in-a-complex-and-rapidly-changing-world-3-27-2020.pdf. Published 2020. Accessed 15th May, 2020.

3. Hansen H, Bieler T, Beyer N, et al. Supervised pulmonary tele-rehabilitation versus pulmonary rehabilitation in severe COPD: a randomised multicentre trial. horax. 2020; 75:413–21.

4. Holland AE, Mahal A, Hill CJ, et al. Home-based rehabilitation for COPD using minimal resources: a randomised, controlled equivalence trial. Thorax. 2017; 72:57–65.

5. Singh SJ, Puhan MA, Andrianopoulos V, et al. An official systematic review of the European Respiratory Society/American Thoracic Society: measurement properties of field walking tests in chronic respiratory disease. Eur Respir J. 2014; 44:1447–78.

6. Brooks D, Solway S, Weinacht K, et al. Comparison between an indoor and an outdoor 6-minute walk test among individuals with chronic obstructive pulmonary disease. Arch Phys Med Rehab. 2003; 84:873–6.

7. Holland AE, Rasekaba T, Fiore JF, Jr., et al. The 6-minute walk distance cannot be accurately assessed at home in people with COPD. Disabil Rehab. 2015; 37:1102–6.

8. Juen J, Cheng Q, Prieto-Centurion V, et al. Health monitors for chronic disease by gait analysis with mobile phones. Telemed J E Health. 2014; 20:1035–41.

9. Juen J, Cheng Q and Schatz B. A natural walking monitor for pulmonary patients using mobile phones. IEEE journal of Biomedical and Health Informatics. 2015; 19:1399–405.

10. Zainuldin R, Mackey MG and Alison JA. Prescription of walking exercise intensity from the 6-minute walk test in people with chronic obstructive pulmonary disease. J Cardiopulm Rehabil Prev. 2015; 35:65–9.

11. Aguilaniu B, Roth H, Gonzalez-Bermejo J, et al. A simple semipaced 3-minute chair rise test for routine exercise tolerance testing in COPD. Int J COPD. 2014; 9:1009–19.

12. Bernabeu-Mora R, Medina-Mirapeix F, Llamazares-Herran E, et al. The accuracy with which the 5 times sit-to-stand test, versus gait speed, can identify poor exercise tolerance in patients with COPD: A cross-sectional study. Medicine. 2016; 95:e4740.

13. Berry MJ, Sheilds KL and Adair NE. Comparison of Effects of Endurance and Strength Training Programs in Patients with COPD. COPD. 2018; 15:192–9.

14. Bossenbroek L, ten Hacken NHT, van der Bij W, et al. Cross-sectional assessment of daily physical activity in chronic obstructive pulmonary disease lung transplant patients. J Heart Lung Transplantation 2009; 28:149–55.

15. Briand J, Behal H, Chenivesse C, et al. The 1-minute sit-to-stand test to detect exercise-induced oxygen desaturation in patients with interstitial lung disease. Ther Adv Resp Dis. 2018; 12:1753466618793028.

16. Butcher SJ, Pikaluk BJ, Chura RL, et al. Associations between isokinetic muscle strength, high-level functional performance, and physiological parameters in patients with chronic obstructive pulmonary disease. Int J COPD. 2012; 7:537–42.

17. Chen Y, Niu Me, Zhang X, et al. Effects of home-based lower limb resistance training on muscle strength and functional status in stable Chronic obstructive pulmonary disease patients. J Clinical Nursing. 2018; 27:e1022–e37.

18. Coquart JB, Le Rouzic O, Racil G, et al. Real-life feasibility and effectiveness of home-based pulmonary rehabilitation in chronic obstructive pulmonary disease requiring medical equipment. Int J COPD. 2017; 12:3549–56.

19. Crook S, Busching G, Schultz K, et al. A multicentre validation of the 1-min sit-to-stand test in patients with COPD. Eur Respir J. 2017; 49.

20. Crook S, Frei A, Ter Riet G, et al. Prediction of long-term clinical outcomes using simple functional exercise performance tests in patients with COPD: a 5-year prospective cohort study. Respir Res. 2017; 18:112.

21. Gloeckl R, Heinzelmann I, Baeuerle S, et al. Effects of whole body vibration in patients with chronic obstructive pulmonary disease--a randomized controlled trial. Respir Med. 2012; 106:75–83.

22. Gonzalez-Saiz L, Fiuza-Luces C, Sanchis-Gomar F, et al. Benefits of skeletal-muscle exercise training in pulmonary arterial hypertension: The WHOLEi+12 trial. Int J Cardiology. 2017; 231:277–283.

23. Grosbois JM, Gicquello A, Langlois C, et al. Long-term evaluation of home-based pulmonary rehabilitation in patients with COPD. Int J COPD. 2015; 10:2037–44.

24. Gruet M, Peyre-Tartaruga LA, Mely L, et al. The 1-Minute Sit-to-Stand Test in Adults With Cystic Fibrosis: Correlations With Cardiopulmonary Exercise Test, 6-Minute Walk Test, and Quadriceps Strength. Respir Care. 2016; 61:1620–8.

25. Hansen H, Beyer N, Frolich A, et al. Intra-and inter-rater reproducibility of the 6-minute walk test and the 30-second sit-to-stand test in patients with severe and very severe COPD. Int J COPD. 2018; 13:3447–57.

26. Jones SE, Kon SSC, Canavan JL, et al. The five-repetition sit-to-stand test as a functional outcome measure in COPD. Thorax. 2013; 68:1015–20.

27. Kongsgaard M, Backer V, Jorgensen K, et al. Heavy resistance training increases muscle size, strength and physical function in elderly male COPD-patients--a pilot study. Respir Med. 2004; 98:1000–7.

28. Levesque J, Antoniadis A, Li PZ, et al. Minimal clinically important difference of 3-minute chair rise test and the DIRECT questionnaire after pulmonary rehabilitation in COPD patients. Int J COPD. 2019; 14:261–9.

29. Li P, Liu J, Lu Y, et al. Effects of long-term home-based Liuzijue exercise combined with clinical guidance in elderly patients with chronic obstructive pulmonary disease. Clin Intervent Aging 2018; 13:1391–9.

30. Mancuso CA, Choi TN, Westermann H, et al. Measuring physical activity in asthma patients: two-minute walk test, repeated chair rise test, and self-reported energy expenditure. J Asthma. 2007; 44:333–40.

31. Morita AA, Bisca GW, Machado FVC, et al. Best Protocol for the Sit-to-Stand Test in Subjects With COPD. Respir Care. 2018; 63:1040–9.

32. Neves CDC, Lacerda ACR, Lage VKS, et al. Whole body vibration training increases physical measures and quality of life without altering inflammatory-oxidative biomarkers in patients with moderate COPD. J App Phys. 2018; 125:520–8.

33. Oliveira A, Afreixo V and Marques A. Enhancing our understanding of the time course of acute exacerbations of COPD managed on an outpatient basis. Int J COPD. 2018; 13:3759–66.

34. Ozalevli S, Ozden A, Itil O, et al. Comparison of the Sit-to-Stand Test with 6 min walk test in patients with chronic obstructive pulmonary disease. Respir Med. 2007; 101:286–93.

35. Puhan MA, Siebeling L, Zoller M, et al. Simple functional performance tests and mortality in COPD. Eur Respir J. 2013; 42:956–63.

36. Radtke T, Hebestreit H, Puhan MA, et al. The 1-min sit-to-stand test in cystic fibrosis - Insights into cardiorespiratory responses. J Cystic Fibrosis. 2017; 16:744–51.

37. Radtke T, Puhan MA, Hebestreit H, et al. The 1-min sit-to-stand test--A simple functional capacity test in cystic fibrosis? J Cystic Fibrosis. 2016; 15:223–6.

38. Reychler G, Boucard E, Peran L, et al. One minute sit-to-stand test is an alternative to 6MWT to measure functional exercise performance in COPD patients. Clin Respir J 2018; 12:1247–56.

39. Regueiro EM, Di Lorenzo VA, Basso RP, et al. Relationship of BODE Index to functional tests in chronic obstructive pulmonary disease. Clinics (Sao Paulo). 2009; 64:983–8.

40. Rietschel E, van Koningsbruggen S, Fricke O, et al. Whole body vibration: a new therapeutic approach to improve muscle function in cystic fibrosis? Int J Rehabilitation Research. 2008; 31:253–6.

41. Rosenbek Minet L, Hansen LW, Pedersen CD, et al. Early telemedicine training and counselling after hospitalization in patients with severe chronic obstructive pulmonary disease: a feasibility study. BMC Med Informatics Decision Making. 2015; 15:3.

42. Sheppard E, Chang K, Cotton J, et al. Functional Tests of Leg Muscle Strength and Power in Adults With Cystic Fibrosis. Respir Care. 2019; 64:40–7.

43. Zanini A, Aiello M, Cherubino F, et al. The one repetition maximum test and the sit-to-stand test in the assessment of a specific pulmonary rehabilitation program on peripheral muscle strength in COPD patients. Int J COPD. 2015; 10:2423–30.

44. Zhang Q, Li Y-X, Li X-L, et al. A comparative study of the five-repetition sit-to-stand test and the 30-second sit-to-stand test to assess exercise tolerance in COPD patients. Int J COPD. 2018; 13:2833–9.

45. Vaidya T, de Bisschop C, Beaumont M, et al. Is the 1-minute sit-to-stand test a good tool for the evaluation of the impact of pulmonary rehabilitation? Determination of the minimal important difference in COPD. Int J COPD. 2016; 11:2609–16.

46. Spielmanns M, Boeselt T, Gloeckl R, et al. Low-Volume Whole-Body Vibration Training Improves Exercise Capacity in Subjects With Mild to Severe COPD. Respir Care. 2017; 62:315–23.

47. Vainshelboim B, Oliveira J, Yehoshua L, et al. Exercise training-based pulmonary rehabilitation program is clinically beneficial for idiopathic pulmonary fibrosis. Respiration 2014; 88:378–88.

48. Benton MJ and Alexander JL. Validation of functional fitness tests as surrogates for strength measurement in frail, older adults with chronic obstructive pulmonary disease. Am J Phys Med Rehab. 2009; 88:579–83; quiz 84-6, 90.

49. Mazzarin C, Kovelis D, Biazim S, et al. Physical Inactivity, Functional Status and Exercise Capacity in COPD Patients Receiving Home-Based Oxygen Therapy. COPD. 2018; 15:271–6.

50. Aurora P, Prasad SA, Balfour-Lynn IM, et al. Exercise tolerance in children with cystic fibrosis undergoing lung transplantation assessment. Eur Respir J.2001; 18:293–7.

51. Basso RP, Jamami M, Pessoa BV, et al. Assessment of exercise capacity among asthmatic and healthy adolescents. Revista brasileira de fisioterapia. 2010; 14:252–8.

52. Bonnevie T, Allingham M, Prieur G, et al. The six-minute stepper test is related to muscle strength but cannot substitute for the one repetition maximum to prescribe strength training in patients with COPD. Int J COPD. 2019; 14:767–74.

53. Bonnevie T, Gravier F-E, Leboullenger M, et al. Six-minute Stepper Test to Set Pulmonary Rehabilitation Intensity in Patients with COPD - A Retrospective Study. COPD. 2017; 14:293–7.

54. Borel B, Fabre C, Saison S, et al. An original field evaluation test for chronic obstructive pulmonary disease population: the six-minute stepper test. Clinical rehab. 2010; 24:82–93.

55. Borel B, Wilkinson-Maitland CA, Hamilton A, et al. Three-minute constant rate step test for detecting exertional dyspnea relief after bronchodilation in COPD. Int J COPD. 2016; 11:2991–3000.

56. Camargo AA, Lanza FC, Tupinamba T, et al. Reproducibility of step tests in patients with bronchiectasis. Brazilian J Phys Ther. 2013; 17:255–62.

57. Coquart JB, Lemaitre F, Castres I, et al. Reproducibility and Sensitivity of the 6-Minute Stepper Test in Patients with COPD. COPD. 2015; 12:533–8.

58. Cox NS, Alison JA, Button BM, et al. Assessing exercise capacity using telehealth: a feasibility study in adults with cystic fibrosis. Respir Care. 2013; 58:286–90.

59. da Costa JN, Arcuri JF, Goncalves IL, et al. Reproducibility of cadence-free 6-minute step test in subjects with COPD. Respir Care. 2014; 59:538–42.

60. Dal Corso S, de Camargo AA, Izbicki M, et al. A symptom-limited incremental step test determines maximum physiological responses in patients with chronic obstructive pulmonary disease. Respir Med. 2013; 107:1993–9.

61. Dal Corso S, Duarte SR, Neder JA, et al. A step test to assess exercise-related oxygen desaturation in interstitial lung disease. Eur Respir J. 2007; 29:330–6.

62. de Camargo AA, Justino T, de Andrade CHS, et al. Chester step test in patients with COPD: reliability and correlation with pulmonary function test results. Respir Care. 2011; 56:995–1001.

63. Delourme J, Stervinou-Wemeau L, Salleron J, et al. Six-minute stepper test to assess effort intolerance in interstitial lung diseases. Sarcoidosis, vasculitis, and diffuse lung diseases. 2012; 29:107–12.

64. Fabre C, Chehere B, Bart F, et al. Relationships between heart rate target determined in different exercise testing in COPD patients to prescribed with individualized exercise training. Int J COPD. 2017; 12:1483–9.

65. Fox BD, Langleben D, Hirsch A, et al. Step climbing capacity in patients with pulmonary hypertension. Clin Res Cardiology. 2013; 102:51–61.

66. Grosbois JM, Riquier C, Chehere B, et al. Six-minute stepper test: a valid clinical exercise tolerance test for COPD patients. Int J COPD. 2016; 11:657–63.

67. Holland AE, Rasekaba T, Wilson JW, et al. Desaturation during the 3-minute step test predicts impaired 12-month outcomes in adult patients with cystic fibrosis. Respir Care. 2011; 56:1137–42.

68. Jose A and Dal Corso S. Step Tests Are Safe for Assessing Functional Capacity in Patients Hospitalized With Acute Lung Diseases. J Cardiopulm Rehab Prev. 2016; 36:56–61.

69. Karloh M, Correa KS, Martins LQ, et al. Chester step test: assessment of functional capacity and magnitude of cardiorespiratory response in patients with COPD and healthy subjects. Brazilian J Physical Therapy. 2013; 17:227–35.

70. Marrara KT, Marino DM, Jamami M, et al. Responsiveness of the six-minute step test to a physical training program in patients with COPD. Jornal brasileiro de pneumologia. 2012; 38:579–87.

71. Murphy N, Bell C and Costello RW. Extending a home from hospital care programme for COPD exacerbations to include pulmonary rehabilitation. Respir Med. 2005; 99:1297–302.

72. Narang I, Pike S, Rosenthal M, et al. Three-minute step test to assess exercise capacity in children with cystic fibrosis with mild lung disease. Pediatric Pulmonology. 2003; 35:108–13.

73. Perrault H, Baril J, Henophy S, et al. Paced-walk and step tests to assess exertional dyspnea in COPD. COPD. 2009; 6:330–9.

74. Pessoa BV, Arcuri JF, Labadessa IG, et al. Validity of the six-minute step test of free cadence in patients with chronic obstructive pulmonary disease. Brazilian J Physical Therapy. 2014; 18:228–36.

75. Pichon R, Couturaud F, Mialon P, et al. Responsiveness and Minimally Important Difference of the 6-Minute Stepper Test in Patients with Chronic Obstructive Pulmonary Disease. Respiration. 2016; 91:367–73.

76. Rusanov V, Shitrit D, Fox B, et al. Use of the 15-steps climbing exercise oximetry test in patients with idiopathic pulmonary fibrosis. Respir Med. 2008; 102:1080–8.

77. Shitrit D, Rusanov V, Peled N, et al. The 15-step oximetry test: a reliable tool to identify candidates for lung transplantation among patients with idiopathic pulmonary fibrosis J Heart Lung Transplantation. 2009; 28:328–33.

78. Starobin D, Kramer MR, Yarmolovsky A, et al. Assessment of functional capacity in patients with chronic obstructive pulmonary disease: correlation between cardiopulmonary exercise, 6 minute walk and 15 step exercise oximetry test. Isr Med Assoc J. 2006; 8:460–3.

79. Tancredi G, Quattrucci S, Scalercio F, et al. 3-min step test and treadmill exercise for evaluating exercise-induced asthma. Eur Respir J. 2004; 23:569–74.

80. Chehere B, Bougault V, Gicquello A, et al. Cardiorespiratory Response to Different Exercise Tests in Interstitial Lung Disease. Med Sci Sports Ex. 2016; 48:2345–52.

81. Rammaert B, Leroy S, Cavestri B, et al. Home-based pulmonary rehabilitation in idiopathic pulmonary fibrosis. Rev Mal Respir. 2011; 28:e52–7.

82. Al Haddad MA, John M, Hussain S, et al. Role of the Timed Up and Go Test in Patients With Chronic Obstructive Pulmonary Disease. J Cardiopulm Rehab Prev. 2016; 36:49–55.

83. Albarrati AM, Gale NS, Enright S, et al. A simple and rapid test of physical performance inchronic obstructive pulmonary disease. Int J COPD. 2016; 11:1785–91.

84. Beauchamp MK, Hill K, Goldstein RS, et al. Impairments in balance discriminate fallers from non-fallers in COPD. Respir Med. 2009; 103:1885–91.

85. Butcher SJ, Meshke JM and Sheppard MS. Reductions in functional balance, coordination, and mobility measures among patients with stable chronic obstructive pulmonary disease. J Cardiopulm Rehab Prev. 2004; 24:274–80.

86. Marques A, Cruz J, Quina S, et al. Reliability, Agreement and Minimal Detectable Change of the Timed Up & Go and the 10-Meter Walk Tests in Older Patients with COPD. COPD. 2016; 13:279–87.

87. Mekki M, Paillard T, Sahli S, et al. Effect of adding neuromuscular electrical stimulation training to pulmonary rehabilitation in patients with chronic obstructive pulmonary disease: randomized clinical trial. Clinical Rehab. 2019; 33:195–206.

88. Mesquita R, Janssen DJ, Wouters EF, et al. Within-day test-retest reliability of the Timed Up & Go test in patients with advanced chronic organ failure. Arch Phys Med Rehabil. 2013; 94:2131–8.

89. Mesquita R, Wilke S, Smid DE, et al. Measurement properties of the Timed Up & Go test in patients with COPD. Chron Respir Dis. 2016; 13:344–52.

90. Vainshelboim B, Kramer MR, Myers J, et al. 8-Foot-Up-and-Go Test is Associated with Hospitalizations and Mortality in Idiopathic Pulmonary Fibrosis: A Prospective Pilot Study. Lung. 2019; 197:81–8.

91. Wilke S, Spruit MA, Wouters EF, et al. Determinants of 1-year changes in disease-specific health status in patients with advanced chronic obstructive pulmonary disease: A 1-year observational study. Int J Nurs Pract. 2015; 21:239–48.

92. Holland AE, Spruit MA, Troosters T, et al. An official European Respiratory Society/American Thoracic Society Technical Standard: field walking tests in chronic respiratory disease. Eur Respir J. 2014; 44:1428–46.

93. Bernabeu-Mora R, Medina-Mirapeix F, Llamazares-Herran E, et al. The Short Physical Performance Battery is a discriminative tool for identifying patients with COPD at risk of disability. Int J COPD. 2015; 10:2619–26.

